# Web design and implementation factors associated with missed opportunities to provide testing on GetCheckedOnline, British Columbia’s digital testing service for sexually transmitted and blood-borne infections: 2022 client experience survey findings

**DOI:** 10.1101/2025.05.16.25327747

**Authors:** Ihoghosa Iyamu, Aidan Ablona, Hsiu-Ju Chang, Heather Pedersen, Devon Haag, Catherine Worthington, Daniel Grace, Amy Salmon, Mieke Koehoorn, Mark Gilbert

## Abstract

**Background:** We assessed associations between web-design/implementation factors and missed opportunities to provide testing via GetCheckedOnline and assessed if these associations were modified by sociodemographic factors.

**Methods:** A cross-sectional survey was conducted in November and December 2022 among clients who indicated needing testing when they created accounts between April and October 2022. Web-design (user interface and experience) and implementation (organization of clinical services around the website) factors were independently modelled against missed opportunities (self-reported inability/unwillingness to test despite needing testing at account creation) using multivariable logistic regression. Effect modification by sociodemographic factors were also conducted.

**Results:** Among 572 respondents needing testing at account creation, 183 (32.0%, 95%CI: 28.18-35.99%) experienced missed opportunities. Web-design factors associated with missed opportunities were difficulty using GetCheckedOnline’s website (adjusted odds ratio (aOR) 3.40, 95%CI:1.68-6.87), while implementation factors were difficulty getting to a laboratory (aOR:3.26, 95%CI:1.97-5.41); perceived inadequacy of tests offered through GetCheckedOnline (aOR:1.81, 95%CI:1.11-2.95) and being likely to complete testing if self-sampling was available (aOR:2.12, 95%CI:1.32-3.42). Findings were consistent in sensitivity analyses but concerns about privacy and security of personal information on GetCheckedOnline (aOR:1.93, 95%CI:1.11-3.35) was associated with missed opportunities. Sociodemographic factors modified associations as respondents with annual income < $20,000CAD, not employed full-time, immigrants, men (who did not agree GetCheckedOnline offered all needed tests) and women (who experienced difficulties getting to a laboratory) had higher odds missed opportunities.

**Conclusions:** Simplifying web-design, ensuring optimal client education, and including more laboratory locations and self-sampling as options for testing, could reduce missed opportunities and promote equitable access to GetCheckedOnline.

**Author Summary:** Digital health interventions like GetCheckedOnline aim to improve access to testing for sexually transmitted and blood-borne infections (STBBIs), but barriers related to web design and service implementation can limit their impact. Our study, based on a 2022 client experience survey, examined how these factors contribute to missed opportunities for testing among GetCheckedOnline users in British Columbia, Canada. We found that 32% of users who created accounts intending to test, reported not testing through the service. They reported barriers including difficulty navigating the website, accessing laboratories, and concerns about the adequacy of available tests. Importantly, these barriers varied across sociodemographic groups, with individuals with lower incomes, immigrants, and women facing the greatest challenges. Our findings suggest that simplifying website navigation, expanding laboratory access, and introducing self-sampling options could reduce missed opportunities and improve equitable access to digital STBBI testing. These insights highlight the need for ongoing, data-driven optimizations to ensure digital health services effectively reach those who face the greatest barriers to care.

## Introduction

Digital interventions for sexually transmitted and blood-borne infections (STBBI) testing have been promoted as cost-effective, equitable and accessible alternatives to clinic-based testing[1,2]. These interventions are especially needed given increased STBBI incidence and prevalence rates particularly among historically marginalized and priority populations like gay, bisexual men who have sex with men (GBMSM), and young people who disproportionately experience barriers to clinic-based testing[3,4]. During the COVID-19 pandemic, these interventions gained further prominence by filling health access gaps caused by public health restrictions and improving access for historically marginalized populations, including GBMSM and racialized minority populations[5,6].

A scoping review by our team revealed limited evidence that digital STBBI testing interventions promote equitable testing given uptake along socioeconomic gradients[7]. While factors explaining apparent reinforcement of health access disparities by these interventions are multifaceted, some are amenable to program modifications[8]. For example, adapting digital interfaces and service design for these interventions using real-world data is more feasible than improving digital access. A review of British Columbia’s (BC) digital STBBI testing service - GetCheckedOnline – revealed between 2014 and 2019, 12% and 30% of over 13,000 users only created accounts and filled test requisitions respectively without testing. Overall, this represents 42% of account creators who are unable to test and could constitute missed opportunities to provide testing for interventions like GetCheckedOnline[7,9]. Drop-offs in users’ progression through touchpoints in STBBI testing is common and may inequitably affect people disparately affected by STBBIs.

Studies exploring modifiable factors are limited regarding digital STBBI testing interventions. Our review suggests person-centred design, emphasis on privacy and security, and integration of sexual and reproductive health services can promote equitable testing[7]. Another study optimizing self-sampling packs for STBBIs found users’ beliefs about testing behavior outcomes, information retention and decision making abilities, were facilitators of testing[10]. Adapting interventions’ design (user interface and experience) and implementation (organization of connected clinical services) to reduce users’ testing burden while emphasizing benefits may address these factors[10]. Studies also describe how users’ social characteristics including digital access and literacy determine their digital STBBI testing access[9,11,12]. Yet, most studies have not explored how users’ perceptions of these digital interventions’ design and implementation may influence access outcomes[13].

Therefore, this study assessed associations between users’ perceptions of web-design/implementation factors and missed opportunities to provide STBBI testing on GetCheckedOnline. The study also assessed if identified associations were modified by sociodemographic (health equity) factors. We hypothesized that self-reported negative perceptions of web-design and implementation factors are associated with missed opportunities to provide testing on GetCheckedOnline. We also hypothesized that identified associations are significantly greater (modified) among historically marginalized populations defined by age, income, employment status, gender, race/ethnicity, and immigration status.

## Methods

### Study design, population, and setting

This study was an online survey of users who created an account with GetCheckedOnline.com, a digital service run by the BC Centre for Disease Control (BCCDC) since 2014 to provide low-barrier STBBI testing, especially for people disproportionately facing clinic-based testing barriers. GetCheckedOnline’s model is well-described elsewhere [2,14]. Users visit GetChecheckedOnline.com, create an account using a valid email address; complete a risk assessment questionnaire and generate a laboratory requisition form; go to a local laboratory to submit serology, urine, and/or self-collected throat and rectal swab specimens for chlamydia, gonorrhea, HIV, hepatitis C and/or syphilis testing; and receive results online if negative, or by phone call from public health nurses if positive with appropriate follow-up[15]. Currently, GetCheckedOnline operates in eight urban, suburban, and rural communities in BC, and between July 2022 and June 2023, it facilitated over 27,000 test episodes (completed tests). This study report complies with the STROBE statement for cross-sectional studies (supplementary material).

### Survey instrument and data collection

The survey instrument was created based on previous GetCheckedOnline research[2,6]. The researchers, health authority partners, GetCheckedOnline’s implementers and Sexual Health Advisory Group (people with lived experiences of marginalization and STBBI testing barriers) co-created the survey (supplementary material). Factors associated with test completion were identified through literature reviews and discussions with implementers experienced with users’ challenges on GetCheckedOnline. These factors were categorized using the theoretical domains framework including those amenable to web-design or implementation modifications[16]. The survey was piloted among 10 community members familiar with GetCheckedOnline and modified for clarity and appropriateness using feedback.

All users who created accounts between April and October 2022 (a 6-month period ending 45 days before the survey), and who were aged 16 years or older, able to complete the survey in English and consented to be contacted for evaluation during account creation were invited to the survey. Survey invitations were sent via email on November 22, with reminders sent 2, 4 and 7 days after. A $20 honorarium was offered to participants who completed the survey. Data was collected using an electronic data capture tool called REDCap. Ethics approval was provided by University of British Columbia’s Behavioral Research Ethics Board (#H18-00437). Based on surveys showing 76% of users testing on GetCheckedOnline experienced testing barriers compared with 87% of clients with accounts who did not test, we estimated a minimum sample size of 440 participants to detect a prevalence odds ratio of missed opportunities of 2 using a power of 80% at a 95% confidence level.

### Exposure, outcome, and confounding variables

The outcome variable was a missed opportunity to provide STBBI testing on GetCheckedOnline which we defined as self-reported inability or unwillingness to test within 45 days of account creation despite reporting needing testing at account creation. This timeline was chosen because program data suggested 90% of users who test do so within 38 days of account creation. Explanatory variables were modifiable web-design factors defined as: a) ease of using the GetCheckedOnline website (easy/not easy); b) clarity of instructions for using GetCheckedOnline (clear/not clear); c) appropriateness of the wording of questions on GetCheckedOnline (appropriate/inappropriate); d) use of GetCheckedOnline based on recommendation from care provider, friends, family or other sources (yes/no); and e) concern about privacy and security of private information when using GetCheckedOnline (concerned/not concerned). Explanatory variables also included modifiable implementation factors defined as: a) ease of getting to a laboratory location (easy/not easy); perceived adequacy of STBBI tests available on GetCheckedOnline (adequate/inadequate); and c) likelihood of testing on GetCheckedOnline with online postal self-collection services (OPSS)) (likely/unlikely). Explanatory variables were dichotomized from 5-point Likert scales as response levels had small samples. Potential effect modifier and confounding variables included *measures of social location*: age, gender, sexual identity, race/ethnicity, highest level of education, language most comfortable speaking with healthcare providers (HCP), employment status, immigrant status, health authority (proxy for geographic residence); *measures of material circumstances*: eHeals score (an eight-item digital health literacy scale scored between 8 and 40 with higher scores implying better literacy)[17], access to a primary HCP, access to a usual place of testing, previous STBBI testing delays due to individual, clinic-related and COVID-19 related reasons, *beliefs about digital health technologies*; convenience of GetCheckedOnline compared with HCP, perceived control of testing through GetCheckedOnline, perceived privacy of testing on GetCheckedOnline compared with in-person; and measures of *sexual behaviors and health*; ever diagnosed with STBBI, number of sexual partners in the past 12 months and participation in condomless anal and/or vaginal sex with multiple sex partners in the past 3 months. These variables informed directed acyclic graphs (DAG) created to describe causal models for each exposure (supplemental material).

### Analytic dataset and statistical analyses

Survey participants were included in this analysis if they indicated they needed to test for STBBIs when creating their accounts (Figure 1). Data was summarized using descriptive statistics. Given missingness in the dataset with 29 variables missing between 0.2% and 23.1% of responses (supplemental material), the main analyses was conducted by imputing missing data in 20 datasets using multivariate imputation by chained equations (MICE) under the missing at random assumption[18]. This assumption was plausible considering missingness patterns were explained by other variables. For example, the health authority variable had higher missingness among respondents with lower education levels, and income had higher missingness among respondents identifying as Indigenous, Black, and other people of color. We imputed 20 datasets based on recommendations to reduce variability from the imputation[19].

**Figure 1:**
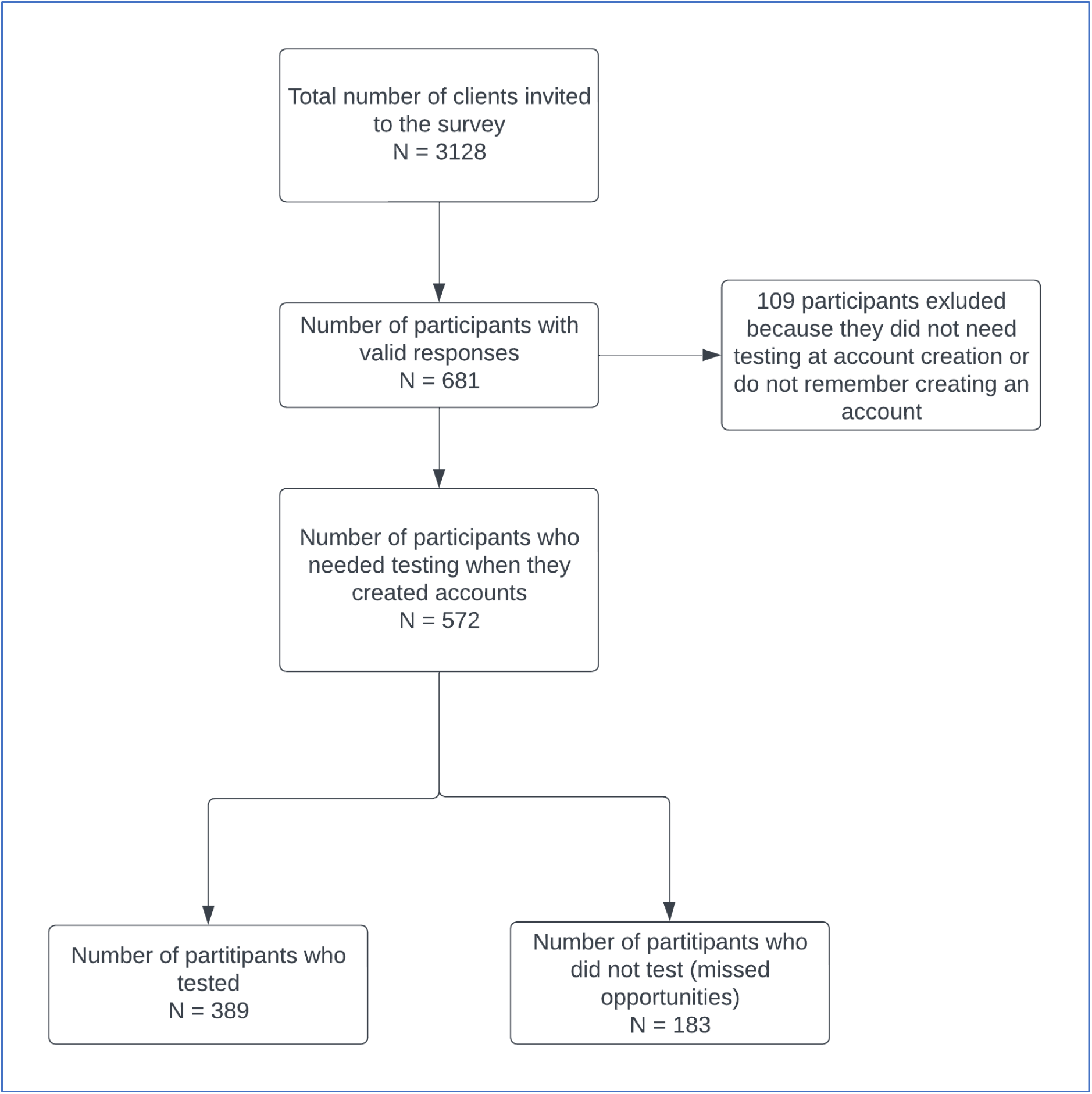
Flow chart of the analytic sample selection using data from the 2022 GetCheckedOnline client experience survey.

Specification of DAGs were guided by the health equity measurement framework which describes the hierarchy of equity factors, suggesting measures of social location (gender, race/ethnicity, and sexual identity, education, income, employment, immigrant status) influence access to material circumstances and digital literacy which in turn determine peoples’ capacity to leverage digital health technologies[20]. The theory of acceptance and use of technology suggests peoples’ expectations of efforts, benefits, social influence and facilitatory conditions of using technology influences intention and utilization, but these influences are modified by age, gender, and experience with the technology[20,21].

Crude and multivariable logistic regression models were run on each imputed dataset using DAGs for variable selection and adjustment based on described theories. Pooled regression estimates and corresponding 95% confidence intervals (95%CI) were extracted using Rubin’s rules[22,23]. Sensitivity analyses were conducted by rerunning models using a complete case dataset (excluding participants with missing data) for each exposure. Each association between web-design/implementation factors and missed opportunities was assessed for effect modification by each sociodemographic factor by testing for significance of effect modification using each measure of social location while adjusting other measures based on the DAGs. Given effect modification analyses were exploratory, the complete-case dataset was used instead of imputed datasets to avoid the computationally complex process of integrating all significant interaction terms in the imputation models which potentially would affect main effect estimates[24]. Estimates were reported from models where effect modification tests suggested a significantly better fit for the data. Each model was assessed for collinearity with variance inflating factor set at <10. Analyses were conducted using R version 3.5.2[25]. Multiple imputation was done using the MICE package[18].

## Results

### Sample characteristics

Among 3128 invited clients, 681 survey responses were received for a response rate of 22%. Among the 681 respondents, 572 (84.0%) indicated needing testing when they created accounts and were included in these analyses (Table 1). Among the 572 included respondents, 177 (30.9%) were aged 30-39 years, 283 (49.5%) identified as women only, 49 (8.6%) identified as gender diverse, 368 (64.3%) had annual incomes of $20,000CAD or more, and 182 (31.8%) resided in the urban city of Vancouver. Overall, 183 (32.0%, 95%CI: 28.18-35.99%) experienced missed opportunities, 57 (10.0%) reported GetCheckedOnline’s website was not easy to use, 53 (9.3%) reported instructions for using GetCheckedOnline were unclear, 37 (6.5%) suggested wording of questions on GetCheckedOnline was inappropriate, 308 (53.8%) were recommended to use GetCheckedOnline by friends/families and other parties, 166 (29.0%) were concerned about privacy and security of personal information when using the website. Further, 135 (23.6%) indicated it was not easy to get to a local laboratory for specimen submission, 134 (23.4%) reported tests offered through GetCheckedOnline were inadequate and 249 (43.5%) said they were more likely to test on GetCheckedOnline if OPSS options are available.

**Table 1:** Characteristics of included participants who needed testing at the time of account creation on GetCheckedOnline - 2022 Client experience survey.

| **Factors** | **Tested on GCO**<br>**N (%)** | **Missed**<br>**opportunity**<br>**N (%)** | **Total**<br>**N (%)** |
| --- | --- | --- | --- |
|  | **389 (68.0)** | **183 (32.0)** | **572 (100.0)** |
| **Sociodemographic factors** |  |  |  |
| **Age** |  |  |  |
| 24 years or less | 78 (20.1) | 35 (19.1) | 113 (19.8) |
| 25-29 years | 97 (24.9) | 28 (15.3) | 125 (21.9) |
| 30-39 years | 114 (29.3) | 63 (34.4) | 177 (30.9) |
| 40 years or more | 56 (14.4) | 29 (15.8) | 85 (14.9) |
| Missing | 44 (11.3) | 28 (15.3) | 72 (12.6) |
| **Health authority** |  |  |  |
| Fraser | 55 (14.1) | 24 (13.1) | 79 (13.8) |
| Interior/Northern | 25 (6.4) | 21 (11.5) | 46 (8.0) |
| Island | 91 (23.4) | 42 (23.0) | 133 (23.3) |
| Vancouver | 136 (35.0) | 46 (25.1) | 182 (31.8) |
| Missing | 82 (21.1) | 50 (27.3) | 132 (23.1) |
| **Race/ethnicity** |  |  |  |
| Indigenous | 19 (4.9) | 6 (3.3) | 25 (4.4) |
| Black and other people of color | 119 (30.6) | 54 (29.5) | 173 (30.2) |
| White | 211 (54.2) | 97 (53.0) | 308 (53.8) |
| Missing | 40 (10.3) | 26 (14.2) | 66 (11.5) |
| **Gender** |  |  |  |
| Gender diverse | 40 (10.3) | 15 (8.2) | 49 (8.6) |
| Man only | 34 (8.7) | 45 (24.6) | 174 (30.4) |
| Woman only | 129 (33.2) | 96 (52.5) | 283 (49.5) |
| Missing | 187 (48.1) | 27 (14.8) | 66 (11.5) |
| **Sexuality** |  |  |  |
| Sexually diverse | 228 (58.6) | 103 (56.3) | 331 (57.9) |
| Straight only | 158 (40.6) | 76 (41.5) | 234 (40.9) |
| Missing | 3 (0.8) | 4 (2.2) | 7 (1.2) |
| **Employed full time** |  |  |  |
| No | 175 (45.0) | 81 (44.3) | 256 (44.8) |
| Yes | 214 (55.0) | 101 (55.2) | 315 (55.1) |
| Missing | 0 (0.0) | 1 (0.5) | 1 (0.2) |
| **Annual income** |  |  |  |
| \$20,000 or more | 256 (65.8) | 112 (61.2) | 368 (64.3) |
| Less than \$20,000 | 69 (17.7) | 34 (18.6) | 103 (18.0) |
| Missing | 64 (16.5) | 37 (20.2) | 101 (17.7) |
| **Immigrant status** |  |  |  |
| Born in Canada | 52 (28.4) | 118 (30.3) | 170 (29.7) |
| Immigrant | 107 (58.5) | 232 (59.6) | 339 (59.3) |
| Missing | 24 (13.1) | 39 (10.0) | 63 (11.0) |
| **Language most comfortable speaking with HCP** |  |  |  |
| English | 324 (83.3) | 153 (83.6) | 477 (83.4) |
| Minority languages | 26 (6.7) | 5 (2.7) | 31 (5.4) |
| Missing | 39 (10.0) | 25 (13.7) | 64 (11.2) |
| **Education** |  |  |  |
| Bachelor's degree or higher | 204 (52.4) | 87 (47.5) | 291 (50.9) |
| Post-secondary education | 82 (21.1) | 40 (21.9) | 122 (21.3) |
| Up to high school | 64 (16.5) | 30 (16.4) | 94 (16.4) |
| Missing | 39 (10.0) | 26 (14.2) | 65 (11.4) |
| **eHeals Score (mean (SD))** | 27.85 (10.59) | 26.87 (11.73) | 27.54 (10.97) |
| **WEB DESIGN FACTORS** |  |  |  |
| **Ease of using GetCheckedOnline Website** |  |  |  |
| Easy | 334 (85.9) | 133 (72.7) | 467 (81.6) |
| Not Easy | 28 (7.2) | 29 (15.8) | 57 (10.0) |
| Missing | 27 (6.9) | 21 (11.5) | 48 (8.4) |
| **Clarity of instructions for using GetCheckedOnline** |  |  |  |
| Not clear | 31 (8.0) | 22 (12.0) | 53 (9.3) |
| Clear | 328 (84.3) | 139 (76.0) | 467 (81.6) |
| Missing | 30 (7.7) | 22 (12.0) | 52 (9.1) |
| **Appropriateness of wording of questions on GetCheckedOnline** |  |  |  |
| Inappropriate | 24 (6.2) | 13 (7.1) | 37 (6.5) |
| Appropriate | 339 (87.1) | 148 (80.9) | 487 (85.1) |
| Missing | 26 (6.7) | 22 (12.0) | 48 (8.4) |
| **Was recommended to use GetCheckedOnline from a friend/known colleague before creating an account** |  |  |  |
| No | 144 (37.0) | 70 (38.3) | 214 (37.4) |
| Yes | 216 (55.5) | 92 (50.3) | 308 (53.8) |
| Missing | 29 (7.5) | 21 (11.5) | 50 (8.7) |
| **Concern about privacy and security of private information when using GetCheckedOnline** |  |  |  |
| Concerned | 108 (27.8) | 58 (31.7) | 166 (29.0) |
| Not concerned | 247 (63.5) | 100 (54.6) | 347 (60.7) |
| Missing | 34 (8.7) | 25 (13.7) | 59 (10.3) |
| **IMPLEMENTATION FACTORS** |  |  |  |
| **Ease of getting to a laboratory location** |  |  |  |
| Easy | 306 (78.7) | 105 (57.4) | 411 (71.9) |
| Not easy | 71 (18.3) | 64 (35.0) | 135 (23.6) |
| Missing | 12 (3.1) | 14 (7.7) | 26 (4.5) |
| **Perceived adequacy of STBBI tests available on GetCheckedOnline** |  |  |  |
| Adequate | 272 (69.9) | 105 (57.4) | 377 (65.9) |
| Inadequate | 80 (20.6) | 54 (29.5) | 134 (23.4) |
| Missing | 37 (9.5) | 24 (13.1) | 61 (10.7) |
| **Likelihood of testing on GetCheckedOnline if online postal self-collection services are available** |  |  |  |
| Likely | 145 (37.3) | 104 (56.8) | 249 (43.5) |
| Unlikely | 216 (55.5) | 57 (31.1) | 273 (47.7) |
| Missing | 28 (7.2) | 22 (12.0) | 50 (8.7) |

GCO: GetCheckedOnline; HCP: Healthcare Provider; SD: Standard Deviation; eHeals: eHealth literacy scale scores; Missed opportunity: defined as self-reported inability or unwillingness to test on GetCheckedOnline despite reporting needing to test at account creation.

### Web-design and implementation factors associated with missed opportunities for STBBI testing

Participants had significantly higher odds of experiencing missed opportunities if they reported GetCheckedOnline’s website was not easy to use (adjusted odds ratio (aOR) 3.14, 95%CI:1.53-6.44), getting to a laboratory location for specimen submission was not easy (aOR:3.26, 95%CI:1.97-5.41), tests offered through GetCheckedOnline were inadequate (aOR:1.81, 95%CI:1.11-2.95), and they were more likely to test on GetCheckedOnline if OPSS options are available (aOR:2.12, 95%CI:1.32-3.42) (Table 2).

**Table 2:**
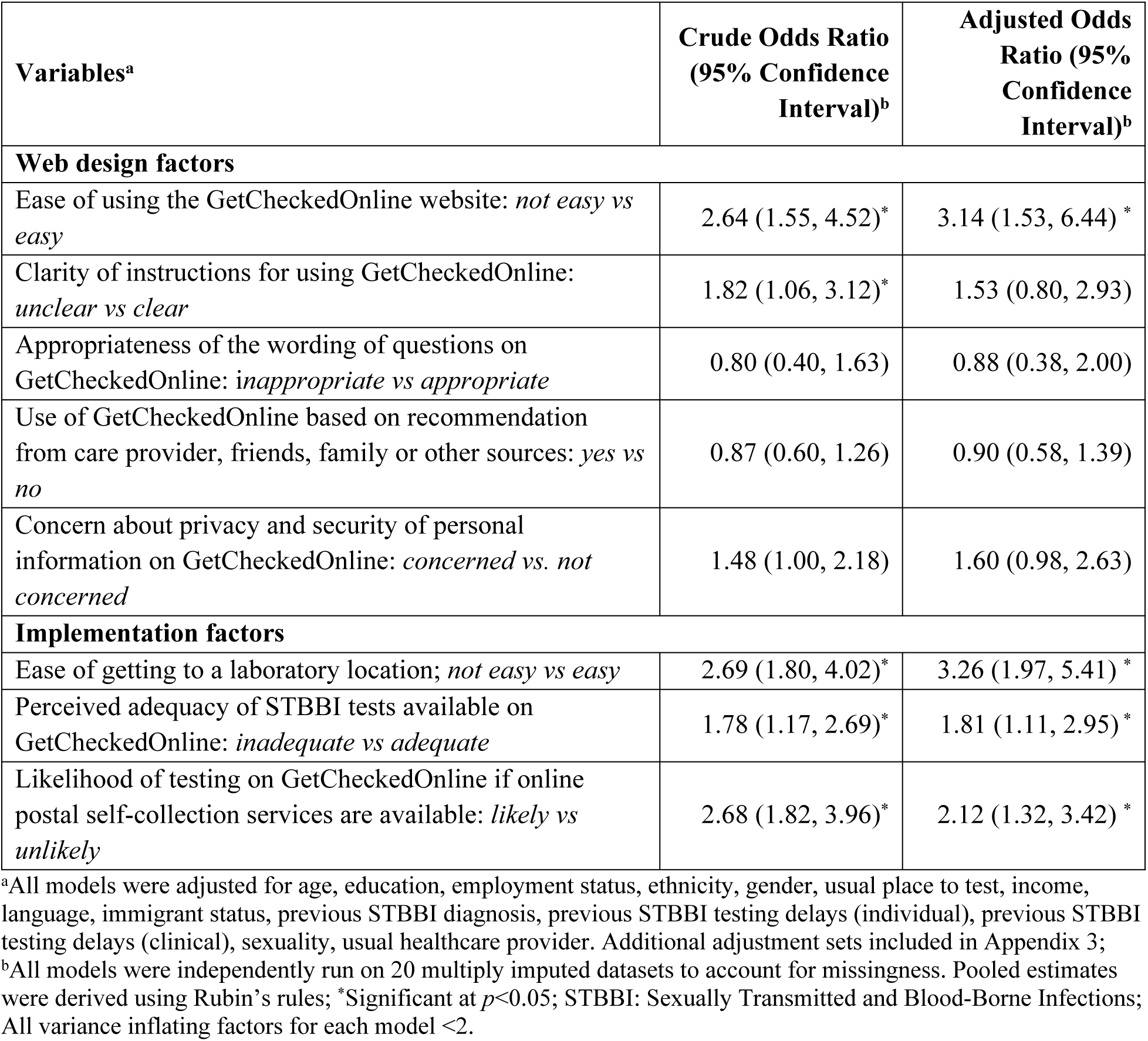
Design and implementation factors associated with missed opportunities for STBBI testing on GetCheckedOnline - 2022 Client experience survey.

| Variables<sup>a</sup> | Crude Odds Ratio (95% Confidence Interval)<sup>b</sup> | Adjusted Odds Ratio (95% Confidence Interval)<sup>b</sup> |
| --- | --- | --- |
| **Web design factors** |  |  |
| Ease of using the GetCheckedOnline website: *not easy vs easy* | 2.64 (1.55, 4.52)* | 3.14 (1.53, 6.44) * |
| Clarity of instructions for using GetCheckedOnline: *unclear vs clear* | 1.82 (1.06, 3.12)* | 1.53 (0.80, 2.93) |
| Appropriateness of the wording of questions on GetCheckedOnline: *inappropriate vs appropriate* | 0.80 (0.40, 1.63) | 0.88 (0.38, 2.00) |
| Use of GetCheckedOnline based on recommendation from care provider, friends, family or other sources: *yes vs no* | 0.87 (0.60, 1.26) | 0.90 (0.58, 1.39) |
| Concern about privacy and security of personal information on GetCheckedOnline: *concerned vs. not concerned* | 1.48 (1.00, 2.18) | 1.60 (0.98, 2.63) |
| **Implementation factors** |  |  |
| Ease of getting to a laboratory location; *not easy vs easy* | 2.69 (1.80, 4.02)* | 3.26 (1.97, 5.41) * |
| Perceived adequacy of STBBI tests available on GetCheckedOnline: *inadequate vs adequate* | 1.78 (1.17, 2.69)* | 1.81 (1.11, 2.95) * |
| Likelihood of testing on GetCheckedOnline if online postal self-collection services are available: *likely vs unlikely* | 2.68 (1.82, 3.96)* | 2.12 (1.32, 3.42) * |

<sup>a</sup>All models were adjusted for age, education, employment status, ethnicity, gender, usual place to test, income, language, immigrant status, previous STBBI diagnosis, previous STBBI testing delays (individual), previous STBBI testing delays (clinical), sexuality, usual healthcare provider. Additional adjustment sets included in Appendix 3;
<sup>b</sup>All models were independently run on 20 multiply imputed datasets to account for missingness. Pooled estimates were derived using Rubin's rules; \*Significant at $p < 0.05$ ; STBBI: Sexually Transmitted and Blood-Borne Infections; All variance inflating factors for each model $< 2$ .

### Effect modification results

Web design and implementation factors showed effect modification with income, employment status, gender, and immigrant status (Table 3). Participants with annual income less than $20,000 CAD who reported GetCheckedOnline’s website was not easy to use had higher odds of experiencing missed opportunities (aOR: 14.95, 95%CI:2.78-80.44). Participants who were not employed full-time and reported instructions on GetCheckedOnline were unclear had higher odds of missed opportunities (aOR: 5.35, 95%CI:1.20-23.96). Participants self-identifying as women only who reported getting to a laboratory was not easy (aOR: 5.60, 95%CI:2.43-12.88), immigrants who reported getting to a laboratory was not easy (aOR: 4.21, 95%CI:2.01-8.81) and participants self-identifying as men only who reported tests available on GetCheckedOnline were inadequate (aOR: 6.95, 95%CI: 2.53-19.10) had higher odds of missed opportunities.

**Table 3:** Design and implementation factors associated with missed opportunities for STBBI testing on GetCheckedOnline, including effect modifiers - 2022 Client experience survey.

| Variables | Adjusted Odds Ratio<br>(95% Confidence Interval) |
| --- | --- |
| **Ease of using the GetCheckedOnline Website and Annual income** |  |
| Annual income (\$20,000 or more): Ease of using the GetCheckedOnline website (not easy vs easy) | 2.28 (0.87, 5.97) |
| Annual income (Less than \$20,000): Ease of using the GetCheckedOnline website (not easy vs easy) | 14.95 (2.78, 80.44)* |
| **Clarity of instructions to use GetCheckedOnline and Employment status** |  |
| Fully employed (No): Clarity of instructions for using GetCheckedOnline (unclear vs clear) | 5.35 (1.20, 23.96)* |
| Fully employed (Yes): Clarity of instructions for using GetCheckedOnline (unclear vs clear) | 0.62 (0.23, 1.67) |
| **Ease of getting to a laboratory to submit specimen and Gender** |  |
| Gender (Gender diverse): Ease of getting to a laboratory location (not easy vs easy) | 0.70 (0.10, 4.93) |
| Gender (Man only): Ease of getting to a laboratory location (not easy vs easy) | 1.23 (0.41, 3.65) |
| Gender (Woman only): Ease of getting to a laboratory location (not easy vs easy) | 5.60 (2.43, 12.88)* |
| **Ease of getting to a laboratory to submit specimen and Immigrant status** |  |
| Immigrant status (Born in Canada): Ease of getting to a laboratory location (not easy vs easy) | 1.00 (0.32, 3.12) |
| Immigrant status (Immigrant): Ease of getting to a laboratory location (not easy vs easy) | 4.21 (2.01, 8.81)* |
| **Perceived adequacy of the tests on GetCheckedOnline and Gender** |  |
| Gender (Gender diverse): Perceived adequacy of STBBI tests available on GetCheckedOnline (inadequate vs adequate) | 1.77 (0.37, 8.52) |
| Gender (Man only): Perceived adequacy of STBBI tests available on GetCheckedOnline (inadequate vs adequate) | 6.95 (2.53, 19.10)* |
| Gender (Woman only): Perceived adequacy of STBBI tests available on GetCheckedOnline (inadequate vs adequate) | 1.30 (0.65, 2.59) |

<sup>a</sup>All models were adjusted for age, gender, sexuality, income, language, education, and race/ethnicity; \*Significant at $p<0.05$ ; STBBI: Sexually Transmitted and Blood-Borne Infections

### Sensitivity analyses

Findings from the sensitivity analyses were consistent with the main findings except for being concerned about privacy and security of personal information on GetCheckedOnline (aOR:1.93, 95%CI:1.11-3.35) which was significantly associated with missed opportunities (Table 4).

**Table 4:** Sensitivity analyses of design and implementation factors associated with missed opportunities on GetCheckedOnline using a complete case dataset (N=304) - 2022 client experience survey.

| **Variables<sup>a</sup>** | **Crude Odds Ratio (95% Confidence Interval)<sup>b</sup>** | **Adjusted Odds Ratio (95% Confidence Interval)<sup>b</sup>** |
| --- | --- | --- |
| **Web design factors** |  |  |
| Ease of using the GetCheckedOnline website: *not easy vs easy* | 2.60 (1.49, 4.54)* | 3.71 (1.61, 8.57)* |
| Clarity of instructions for using GetCheckedOnline: *unclear vs clear* | 1.67 (0.94, 2.99) | 1.19 (0.54, 2.62) |
| Appropriateness of the wording of questions on GetCheckedOnline: *inappropriate vs appropriate* | 0.81 (0.40, 1.63) | 0.78 (0.29, 2.12) |
| Use of GetCheckedOnline based on recommendation from care provider, friends, family or other sources: *yes vs no* | 0.88 (0.60, 1.28) | 0.92 (0.56, 1.50) |
| Concern about privacy and security of personal information on GetCheckedOnline: *concerned vs. not concerned* | 1.33 (0.89, 1.97) | 1.93 (1.11, 3.35)* |
| **Implementation factors** |  |  |
| Ease of getting to a laboratory location; *not easy vs easy* | 2.63 (1.75, 3.93)* | 2.77 (1.50, 5.13)* |
| Perceived adequacy of STBBI tests available on GetCheckedOnline: *inadequate vs adequate* | 1.75 (1.16, 2.64)* | 2.16 (1.27, 3.68)* |
| Likelihood of testing on GetCheckedOnline if online postal self-collection services are available: *likely vs unlikely* | 2.72 (1.85, 4.00)* | 1.80 (1.05, 3.09)* |

<sup>a</sup>All models were adjusted for age, gender, sexuality, income, language, education, and race/ethnicity; <sup>b</sup>All models were independently run on a complete case dataset (N=304); \*Significant at $p < 0.05$ ; STBBI: Sexually Transmitted and Blood-Borne Infections; All variance inflating factors for each model $< 2$ .

## Discussion

Overall, 32% of survey participants experienced missed opportunities to be provided STBBI testing on GetCheckedOnline. The web design factor - ease of use of GetCheckedOnline’s website was associated with higher odds of missed opportunities. Implementation factors, including difficulty accessing a laboratory location for specimen submission, perceived inadequacy of STBBI tests offered on GetCheckedOnline, and greater likelihood of use if self-collection (i.e., OPSS) is available, were also associated with missed opportunities. Sensitivity analyses suggested another web design factor - participants’ concerns about privacy and security of information when using GetCheckedOnline may also be associated with missed opportunities. We found variations in these associations by sociodemographic factors. Users with lower annual income who reported GetCheckedOnline’s website was not easy to use, users not employed full-time who reported instructions on GetCheckedOnline were unclear, self-identified women and immigrants who indicated getting to a laboratory was not easy and self-identified men who reported tests offered through GetCheckedOnline were inadequate had higher odds of missed opportunities.

This study’s findings are congruent with studies of OPSS services that suggest 18% and 42% of users are unable to return test kits after creating accounts and making test requisitions[9,26]. Prior studies suggest person-centred web design and emphasis on privacy are associated with equitable uptake of digital STBBI testing[7,27]. These studies did not directly assess the relationship between web-design and implementation factors and completing STBBI testing as done in our study. This study extends the literature by highlighting the role of ease of website use and concerns about privacy in facilitating digital STBBI testing. It also emphasizes the greater role implementation factors play, even beyond web-design factors. Moreover, while other studies have not explicitly assessed these factors with an equity lens, this study highlights the importance of web-design and implementation factors among low-income, non-fully employed, immigrant users. Findings also highlight potential differences in the impact of these factors based on social factors as proposed by the HEMF[20]. Further, it suggests that women’s experience of inequities in accessing laboratory locations may impact experiences of missed opportunities while men may experience missed opportunities if STBBI tests online are considered inadequate. This may be explained by the population of GBMSM who have historically been better informed about STBBI’s and may consider additional tests essential to their testing experience[28].

The findings demonstrate gaps in uptake of digital STBBI testing despite GetCheckedOnline’s person-centred approaches and focus on equity[14]. Our findings suggest that ensuring easier web navigation/use through simplified web interfaces and further promoting a sense of privacy of personal information on the website can reduce missed opportunities to provide testing on GetCheckedOnline. Given identified associations, providing additional laboratory locations and testing options including self-sample collection can reduce missed opportunities and address access disparities. OPSS and other similar self-collection and self-testing services may improve access especially for people not well served by GetCheckedOnline’s current model, including people who live and work far from existing laboratory locations (e.g., rural, and remote areas). Such options could facilitate access to digital STBBI testing among other historically marginalized groups like women, immigrants and low income earners who may need resources for testing[29]. For example, accessing laboratory locations requires additional flexible resources including paid time off work and transportation[30]. Women and immigrants historically experience work-related and pay disparities that reduce access to flexible resources required to leverage GetCheckedOnline’s current model. Providing laboratory options closer to users or through OPSS will reduce the perceived effort required for testing behaviours in such groups[29] and improve access[21].

However, to inform service modifications, we must further understand users’ experiences of missed opportunities using qualitative approaches. This is especially true among historically marginalized groups with experiences that are not entirely captured through surveys. For example, what aspects of GetCheckedOnline’s website are difficult to use, or what user contexts explain difficulties getting to the laboratory or eagerness to use alternative options like OPSS? This is the subject of ongoing work for this group to update the website and consider additional service options to promote testing access especially among groups experiencing the most barriers with our current design [31].

### Strengths and limitations

This is one of the first studies exploring design and implementation factors associated with missed opportunities to provide digital STBBI testing using theory informed regression. Our analyses account for missing data to address potential non-response biases within the dataset and the consistency of our findings between the main and sensitivity analyses reassures us of the accuracy of our estimates.

However, there are limitations. First, missed opportunities were defined from service providers’ perspectives which are not always aligned with users’ perspectives. The outcome was assigned irrespective of testing recommendations. Further studies are needed to describe missed opportunities among users meeting recommendations for digital STBBI testing but not testing. This study simplified missed opportunities as one single behavior as against a multistep behavior including creating laboratory requisition, accessing laboratories, and completing testing. Multistep behaviours were not explored given sample size limitations. Our simplified approach may conceal relative effects of web-design factors which are more relevant for laboratory requisition. However, studies suggest laboratory requisition is a smaller barrier compared with specimen submission[9,26]. Our use of multiple imputation may have yielded conservative estimates of observed associations due to increased standard errors. Given this was a cross-sectional study, causality cannot be fully established. Further, using self-reported measures increased risks for social desirability and recall bias which may have biased associations towards the null. However, given the survey was online and anonymous, it is unlikely this is significant. Given response rates of 22%, there may be selection bias especially based on varied time between account creation and participation, alongside sociodemographic factors. However, comparing invitees’ and respondents’ characteristics suggests this may be partly addressed with our missingness analyses (supplementary material). Our findings are specific to GetCheckedOnline but may be cautiously generalized to digital STBBI testing models that do not require seeing an HCP for test requisition but require visits to laboratories for specimen submission.

### Conclusion

We found associations between missed opportunities to provide STBBI testing on GetCheckedOnline and web-design and implementation factors. The associations were more apparent for marginalized groups like income earners, women, and immigrants, highlighting the need for ongoing service adaptations. Simplifying the web-interface, emphasizing measures to protect privacy of sensitive information, and providing information about the adequacy of tests available on GetCheckedOnline may reduce missed opportunities while improving equitable access. Further, creating options for self-sample collection can improve access for people experiencing difficulties accessing local laboratory locations, especially for those in rural and remote locations within BC where laboratories are not readily accessible and for people experiencing difficulties accessing local laboratories due to socioeconomic barriers. Finally, our study demonstrates the utility of implementation science studies in optimizing digital STBBI testing systems by using data to drive person-centred optimizations for equity.

## Acknowledgements

We would like to thank GetCheckedOnline users who generously shared their time and experiences with us. We would also like to thank members of the BCCDC’s Clinical Prevention Services – Sexual Health Advisory Group who reviewed study findings and informed our interpretation of the data.

## Declarations

### Ethical approval

Ethics approval was obtained from the University of British Columbia’s Behavioral Ethics Board (ethics #H18-00437).

### Guarantor

Mark Gilbert

### Author Contributions

- Conceptualization: Ihoghosa Iyamu (II), Aidan Ablona (AA), Mark Gilbert (MG)
- Methodology: Ihoghosa Iyamu (II), Aidan Ablona (AA), Hsiu-Ju Chang (HJC), Mark Gilbert (MG)
- Formal Analysis: Ihoghosa Iyamu (II)
- Investigation: Ihoghosa Iyamu (II), Aidan Ablona (AA), Hsiu-Ju Chang (HJC)
- Data Curation: Ihoghosa Iyamu (II), Hsiu-Ju Chang (HJC), Aidan Ablona (AA)
- Writing – Original Draft Preparation: Ihoghosa Iyamu (II)
- Writing – Review & Editing: Ihoghosa Iyamu (II), Hsiu-Ju Chang (HJC), Heather Pedersen (HP), Devon Haag (DH), Catherine Worthington (CW), Daniel Grace (DG), Mark Gilbert (MG)
- Visualization: Ihoghosa Iyamu (II)
- Supervision: Mark Gilbert (MG), Amy Salmon (AS), Mieke Koehoorn (MK)
- Project Administration: Ihoghosa Iyamu (II), Hsiu-Ju Chang (HJC)
- Funding Acquisition: Mark Gilbert (MG), Catherine Worthington (CW), Daniel Grace (DG) All authors have read and approved the final manuscript.

### Data availability statement

The data underlying this study are part of ongoing program evaluation work at the BC Centre for Disease Control (BCCDC). While these data are not publicly available, they can be provided upon reasonable request, subject to institutional approvals and data sharing policies. Requests for access should be directed to the corresponding author, Ihoghosa Iyamu, or the BCCDC’s data governance committee.

## References

1 Exten C, Pinto CN, Gaynor AM, et al. Direct-to-Consumer STI Testing Services: A Position Statement from the American Sexually Transmitted Diseases Association. Sexually transmitted diseases. Published Online First: 2021. doi: 10.1097/OLQ.0000000000001475

2 Gilbert M, Thomson K, Salway T, et al. Differences in experiences of barriers to STI testing between clients of the internet-based diagnostic testing service GetCheckedOnline.com and an STI clinic in Vancouver, Canada. Sexually Transmitted Infections. 2019;95:151–6. doi: 10.1136/sextrans-2017-053325

3 Public Health Agency of Canada. Reducing the health impact of sexually transmitted and blood-borne infections in Canada by 2030: A pan-Canadian STBBI framework for action. Ottawa, Canada: Public Health Agency of Canada 2018.

4 Public Health Agency of Canada Government of Canada. Report on sexually transmitted infections in Canada: 2019. Ottawa, Canada: Public Health Agency of Canada 2021.

5 Gilbert M, Chang H-J, Ablona A, et al. Accessing needed sexual health services during the COVID-19 pandemic in British Columbia, Canada: a survey of sexual health service clients. Sex Transm Infect. 2022;98:360–5. doi: 10.1136/sextrans-2021-055013

6 Iyamu I, Pedersen H, Ablona A, et al. Evaluating the Impact of the COVID-19–Related Public Health Restrictions on Access to Digital Sexually Transmitted and Blood-Borne Infection Testing in British Columbia, Canada: An Interrupted Time Series Analysis. Sexual Trans Dis. 2023;50:595–602. doi: 10.1097/OLQ.0000000000001833

7 Iyamu I, Sierra-Rosales R, Estcourt CS, et al. Differential uptake and effects of digital sexually transmitted and bloodborne infection testing interventions among equity-seeking groups: a scoping review. Sex Transm Infect. 2023;sextrans-2023-055749. doi: 10.1136/sextrans-2023-055749

8 Gómez-Ramírez O, Iyamu I, Ablona A, et al. On the imperative of thinking through the ethical, health equity, and social justice possibilities and limits of digital technologies in public health. Canadian Journal of Public Health. 2021;112:412–6. doi: 10.17269/s41997-021-00487-7

9 Manavi K, Hodson J. Observational study of factors associated with return of home sampling kits for sexually transmitted infections requested online in the UK. BMJ open. 2017;7:e017978. doi: 10.1136/bmjopen-2017-017978

10 Flowers P, Vojt G, Pothoulaki M, et al. Using the behaviour change wheel approach to optimize self-sampling packs for sexually transmitted infection and blood borne viruses. British Journal of Health Psychology. 2022;n/a. doi: 10.1111/bjhp.12607

11 Barnard S, Free C, Bakolis I, et al. Comparing the characteristics of users of an online service for STI self-sampling with clinic service users: a cross-sectional analysis. Sexually transmitted infections. 2018;94:377–83. doi: 10.1136/sextrans-2017-053302

12 Crawford A, Serhal E. Digital Health Equity and COVID-19: The Innovation Curve Cannot Reinforce the Social Gradient of Health. Journal of medical Internet research. 2020;22:e19361. doi: 10.2196/19361

13 Eruchalu CN, Pichardo MS, Bharadwaj M, et al. The Expanding Digital Divide: Digital Health Access Inequities during the COVID-19 Pandemic in New York City. Journal of Urban Health. 2021;98:183–6. doi: 10.1007/s11524-020-00508-9

14 Gilbert M, Haag D, Hottes TS, et al. Get Checked…Where? The Development of a Comprehensive, Integrated Internet-Based Testing Program for Sexually Transmitted and Blood-Borne Infections in British Columbia, Canada. JMIR research protocols. 2016;5:e186. doi: 10.2196/resprot.6293

15 Gilbert M, Salway T, Haag D, et al. Use of GetCheckedOnline, a Comprehensive Web-based Testing Service for Sexually Transmitted and Blood-Borne Infections. Journal of medical Internet research. 2017;19:e81. doi: 10.2196/jmir.7097

16 Atkins L, Francis J, Islam R, et al. A guide to using the Theoretical Domains Framework of behaviour change to investigate implementation problems. Implementation Science. 2017;12:77. doi: 10.1186/s13012-017-0605-9

17 Norman CD, Skinner HA. eHEALS: The eHealth Literacy Scale. J Med Internet Res. 2006;8:e27. doi: 10.2196/jmir.8.4.e27

18 van Buren S, Groothuis-Oudshoorn K. Multivariate Imputation by Chained Equations in R. Journal of Statistical Software. 2011;45:1–67.

19 Sterne JAC, White IR, Carlin JB, et al. Multiple imputation for missing data in epidemiological and clinical research: Potential and pitfalls. BMJ. 2009;338:b2393. doi: 10.1136/bmj.b2393

20 Dover DC, Belon AP. The health equity measurement framework: a comprehensive model to measure social inequities in health. International Journal for Equity in Health. 2019;18:36. doi: 10.1186/s12939-019-0935-0

21 Venkatesh V, Thong JYL, Xu X. Consumer Acceptance and Use of Information Technology: Extending the Unified Theory of Acceptance and Use of Technology. MIS Quarterly. 2012;36:157–78. doi: 10.2307/41410412

22 Lumley T. Complex Surveys: A Guide to Analysis Using R: A Guide to analysis Using R. John Wiley and Sons Inc. 2010.

23 Rubin D. Multiple Imputation for Nonresponse in Surveys. John Wiley and Sons Inc. 1987.

24 Enders CK. Applied missing data analysis. 2nd ed. New York: The Guilford Press 2022.

25 R Core Team. R: A language and environment for statistical computing. 2021.

26 Ricca AV, Hall EW, Khosropour CM, et al. Factors Associated with Returning At-Home Specimen Collection Kits for HIV Testing among Internet-Using Men Who Have Sex with Men. Journal of the International Association of Providers of AIDS Care. 2016;15:463–9.

27 van Bergen JEAM, Fennema JSA, van den Broek IVF, et al. Rationale, design, and results of the first screening round of a comprehensive, register-based, Chlamydia screening implementation programme in the Netherlands. BMC infectious diseases. 2010;10:293. doi: 10.1186/1471-2334-10-293

28 Knight R, Falasinnu T, Oliffe JL, et al. Integrating gender and sex to unpack trends in sexually transmitted infection surveillance data in British Columbia, Canada: an ethno-epidemiological study. BMJ Open. 2016;6:e011209. doi: 10.1136/bmjopen-2016-011209

29 Venkatesh V, Morris MG, Davis GB, et al. User Acceptance of Information Technology: Toward a Unified View. MIS Quarterly. 2003;27:425–78. doi: 10.2307/30036540

30 Phelan JC, Link BG, Tehranifar P. Social Conditions as Fundamental Causes of Health Inequalities: Theory, Evidence, and Policy Implications. Journal of Health and Social Behavior. 2010;51:S28–40.

31 Iyamu I, Kassam R, Worthington C, et al. Missed opportunities to provide sexually transmitted and blood-borne infections testing in British Columbia: An interpretive description of users’ experiences of Get Checked Online’s design and implementation. Digital Health. 2024;10:20552076241277653. doi: 10.1177/20552076241277653

